# Maturity of brain structures and white matter connectomes, and their relationship with psychiatric symptoms in youth

**DOI:** 10.1101/2020.03.02.20029488

**Authors:** Alex Luna, Joel Bernanke, Jiook Cha, Jonathan Posner

**Affiliations:** Department of Psychiatry, Columbia University College of Physicians and Surgeons; Data Science Institute, Columbia University; Department of Psychology, Seoul National University, Seoul, South Korea; New York State Psychiatric Institute

**Keywords:** Machine Learning, Connectome, Diffusion Tensor Imaging, Biomarkers, Psychopathology, brain age

## Abstract

**Background:** Brain neuromaturation can be indexed using brain predicted age difference (BrainPAD), a metric derived by the application of machine learning (ML) algorithms to neuroimaging. Previous studies in youth have been limited to a single type of imaging data, single ML approach, or specific psychiatric condition. Here, we use multimodal neuroimaging and an ensemble ML algorithm to estimate BrainPAD and examine its relationship with broad measures of symptoms and functioning in youth.

**Methods:** We used neuroimaging from eligible participants in the Healthy Brain Network (HBN, N = 498). Participants with a Child Behavior Checklist Total Problem T-Score < 60 were split into training (N=215) and test sets (N=48). Morphometry estimates (from structural MRI), white matter connectomes (from diffusion MRI), or both were fed to an automated ML pipeline to develop BrainPAD models. The most accurate model was applied to a held-out evaluation set (N=249), and the association with several psychometrics was estimated.

**Results:** Models using morphometry and connectomes together had a mean absolute error of 1.16 years, outperforming unimodal models. After dividing participants into positive, normal, and negative BrainPAD groups, negative BrainPAD values were associated with more symptoms on the Child Behavior Checklist (negative=71.6, normal 59.0, p=0.011) and lower functioning on the Children’s Global Assessment Scale (negative=49.3, normal=58.3, p=0.002). Higher scores were associated with better performance on the Flanker task (positive=62.4, normal=52.5, p=0.006).

**Conclusion:** These findings suggest that a multimodal approach, in combination with an ensemble method, yields a robust biomarker correlated with clinically relevant measures in youth.

## Introduction

Human neuromaturation is the complex process that governs the formation and refinement of the structures and connections of the central nervous system from conception through early adulthood. Whereas the mechanisms underlying neuromaturation – such as neural migration, myelination, and synaptic pruning – are relatively conserved across individuals (1), the rates at which these processes unfold are heterogeneous (2). Deviations in the rate of neuromaturation can lead to differences in brain structure, connectivity, and function, with potential implications for the etiology and phenomenology of cognitive impairments and psychopathology (3). These links, however, between neuromaturation, cognition, and psychiatric outcomes remain relatively unexplored, particularly in youth.

Peak incidence for most major psychiatric disorders including depression, anxiety disorders, schizophrenia, and substance use disorders all occur during adolescence, the same period that neuromaturation is at an apex (4). Altered rates of neuromaturation during this period can be indexed by deviations between an individual’s chronological age relative to his or her “brain age” – giving rise to “neuroimmaturity” or “neurosupermaturity.” Brain age and its utility as a marker of disease has been examined in other areas of medicine, including Alzheimer’s disease (5, 6), multiple sclerosis (7, 8), Down’s syndrome (9), and overall mortality (10). To our knowledge, however, few studies have examined the contribution of neuroimmaturity and neurosupermaturity, as measured by estimations of brain age from different types of neuroimaging data, to psychopathology, but preliminary work applying machine learning (ML) algorithms to neuroimaging data supports this approach (11-13).

For example, among studies utilizing structural imaging data, one study noted that individuals with schizophrenia had “older” brains relative to healthy controls (14), while another found that teenage participants at high-risk for psychosis with overestimated brain ages were more likely to become psychotic than participants with “normal” brain ages (15). Functional patterns characteristic of “older” and “younger” states have also been linked to negative outcomes. Namely, “younger” functional states have been associated with increased risk-taking behavior (16). However, the association between differences in brain age and dysfunction is not always clear: neurosupermaturity has also been linked to increased processing speed (17).

Previous tools have examined brain morphology and functional imaging data, but only a select few have incorporated white matter connectomes, much less brain morphology and white matter connectomes together (17, 18). Studies using MRI-derived whole-brain connectomes suggest that atypical development of the connectome is associated with long-term difficulties with emotion, cognition, and behavior in infants and adolescents (19, 20). For example, white matter connectomes at birth were predictive of cognitive performance at age 2 in both full-term and preterm infants (21). Among adolescents with attentional problems, temporoparietal connections were found to have lower fractional anisotropy (22). Thus, the inclusion of white matter connectomes is likely important to studies characterizing the relationship between brain development and psychiatric symptoms, functioning, and cognition in youth (23).

The choice of analytic method has implications for the accuracy and generalizability of the resulting model (24). Previous studies using ML to predict brain age have often employed a single ML algorithm, such as Support Vector Regression (14, 24), Relevance Vector Regression (24), or convolutional neural networks (25). Employing a pipeline that systematically tests multiple preprocessing strategies and ML algorithms, and chooses the most accurate one, could improve accuracy (26). In addition, ensemble learning that combines several individual ML algorithms could also improve model performance (27).

Here, we use an automated ML pipeline that incorporates multiple ML algorithms, including an ensemble method, to estimate brain age from morphometry estimates and whole-brain white matter connectomes obtained from participants in the Healthy Brain Network (HBN), a large community-based cohort study of children and adolescents. We then apply the best ML model to a held-out evaluation dataset of participants with and without a high likelihood for psychopathology to identify associations between deviations in brain age, difficulties with cognitive functioning, and psychiatric symptoms. To our knowledge, this is the first study to use morphometry, whole-brain connectomes, and an ensemble ML algorithm to look at broad measures of symptoms and functioning in children and adolescents.

## Methods and Materials

### Data Source

We used neuroimaging and psychometric data from the HBN collected between 2015, when the study was initiated, through 2017, when HBN neuroimaging data was made publicly available. All phenotypic information was obtained in accordance with the Data Usage Agreement required by the HBN. Briefly, the HBN recruited a community-based sample of healthy and non-healthy boys and girls between ages 5 and 21 in multiple sites in New York City. Participants were excluded from the HBN if they had immediate safety concerns, medical conditions that would confound neuroimaging research, or their symptoms interfered with the participation in the study. Detailed inclusion and exclusion criteria are available elsewhere (28). The HBN study was approved by the Chesapeake Institutional Review Board Written consent from their legal guardians and written assent were obtained from the participants.

### Brain Morphometry

FreeSurfer 6.0 (https://surfer.nmr.mgh.harvard.edu/) was used to generate 678 morphometric estimates from structural magnetic resonance imaging (sMRI) data, including thickness, volumes, surface, and mean curvature, for each participant. The Desikan-Killiany atlas was used (29). In short, structural image processing with FreeSurfer included motion correction, removal of non-brain tissue, Talairach transformation, segmentation, intensity normalization, tessellation of the gray matter/white matter boundary, topology correction, and surface deformation. Deformation procedures used both intensity and continuity information to produce representation of cortical thickness. The maps produced were not restricted to the voxel resolution of the original scans and were thus capable of detecting submillimeter differences. Cortical thickness measures have been validated against histological analysis and manual measurements (30, 31). Further information about this process is provided elsewhere (29, 31-33).

### White Matter Connectomes

To obtain accurate brain phenotypic estimates, we used an individualized connectome approach, rather than population-based regions of interest or tracts of interest methods. We used the Mrtrix (34) MRI analysis pipeline to preprocess diffusion MRI (dMRI), estimate whole-brain white matter tracts, and generate individualized connectome features. Generated features included number of estimated streamlines within a given connection, a commonly used measure of fiber connection strength (35, 36), and fractional anisotropy (FA) from the diffusion tensor model. dMRI images were de-noised (37), motion corrected (38), and then processed through the Advanced Normalization Tools (ANTs) pipeline using the N4 algorithm (39). Probabilistic tractography was performed using 2^nd^-order integration over fiber orientation distributions with a whole-brain streamline count of 20 million (40).

To discard potential false positive streamlines and improve biological plausibility, initial tractograms were filtered using spherical-deconvolution informed filtering with a final streamline count target of 10 million (2:1 ratio). Using the filtered tractogram, an 84 x 84 whole-brain connectome matrix was generated for each participant using the T1-based parcellation and segmentation from FreeSurfer, which was then registered and warped to the participant’s diffusion MRI (b0 images) using ANTs. With this approach, each participant’s white matter connectome estimates were constrained by his or her own neuroanatomy. A total of 7,140 connectome estimates were obtained, weighted by streamline count and FA. Computation was done on supercomputers at Argonne Leadership Computing Facility Theta and Texas Advanced Computing Center Stampede2.

### Brain age model development and comparison

We first divided the HBN dataset into two groups based on the Child Behavior Checklist (CBCL) Total Problem T-Score. The CBCL is a well validated, parent report measure of emotional, behavioral, and social difficulties in children and adolescents, and Total Problem T-scores of 60 or greater are associated with an increased likelihood of psychopathology (41). To train our brain age prediction on a group of likely typically developing children, we used a cutoff of 60, such that participants scoring 60 or above were in one group and below 60 in another. We then further divided the group with CBCL Total Problem T-scores below 60 into a ***training dataset*** *(n=215)*, to develop the age prediction models, and a ***testing dataset*** *(n=48)*, to assess their accuracy, using a random 80%/20% split. Lastly, we created an ***evaluation dataset*** *(n=249)* that consisted of participants with a high likelihood for psychopathology (those with CBCL Total Problem T-Scores of 60 or greater) and a low likelihood for psychopathology (participants from the testing dataset, all of whom had CBCL Total Problem T-Scores less than 60). By pooling participants, the evaluation data set was designed to show that any observed relationships between brain age and outcome measures were robust across the full range of CBCL T scores. Participants without age, sex, race, or ethnicity data were excluded from the evaluation dataset (Figure 1).

**Figure 1:**
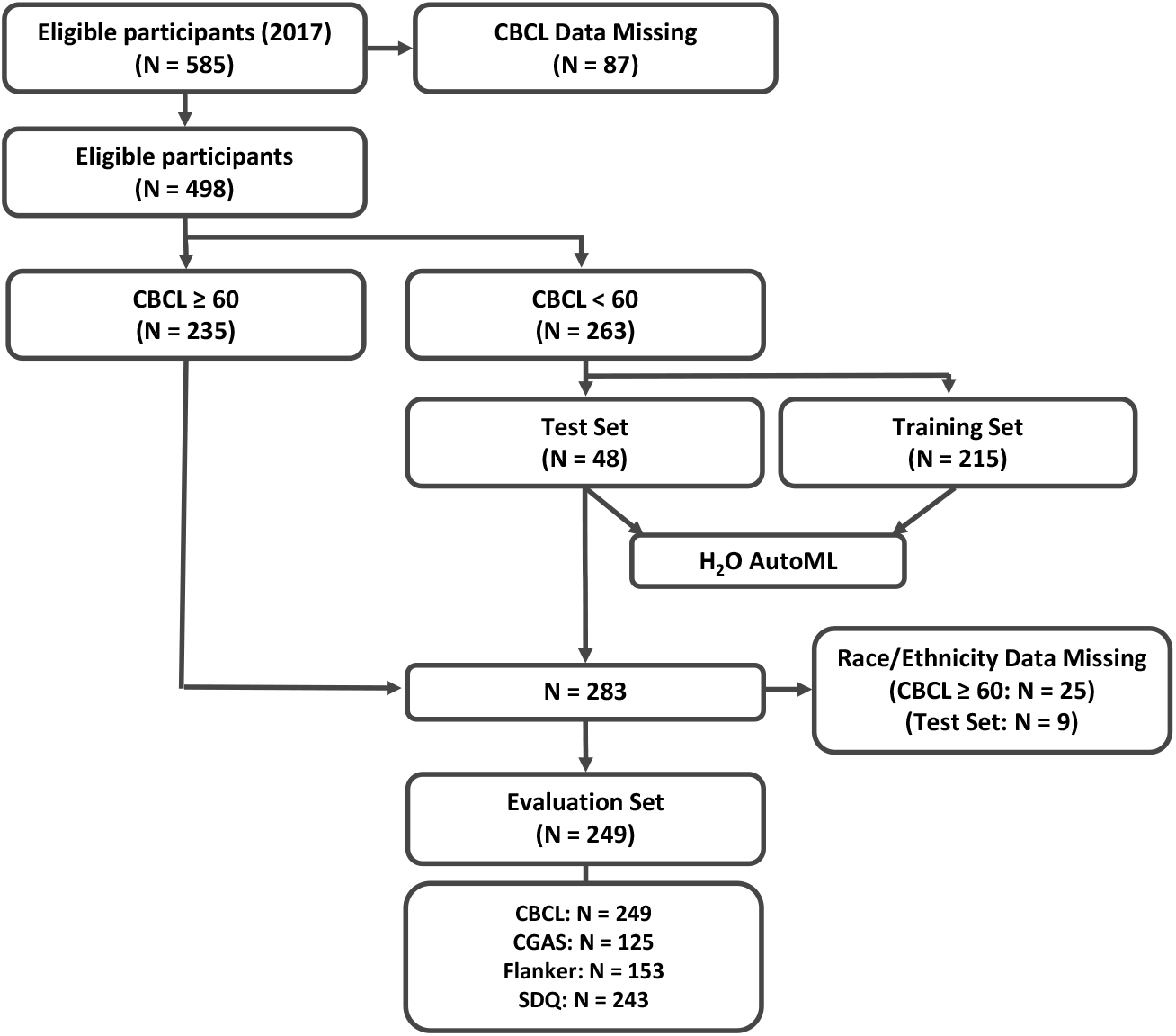
Participant Selection Workflow. In brief, of 498 participants with CBCL-Parent report data available, 263 with a CBCL score <60 were used for modeling with H2O’s AutoML function using an 80%/20% train/test split. For statistical analysis, test set participants and participants with CBCL >59 with race/ethnicity data available were combined for a total of 249 participants. For each metric assessed, participants were split into Negative, Positive, and Normal BrainPAD groups.

We used H_2_O AutoML, an open-source automatic ML pipeline (http://docs.h2o.ai/h2o/latest-stable/h2o-docs/automl.html), to develop and select three brain age prediction models, one for each MRI modality – morphometry (***sMRI alone***) and connectomes (***dMRI alone***) – and a third based on both modalities combined (***sMRI and dMRI combined***) (42). H_2_O AutoML features multiple ML algorithms and performs scalable, automated model training and hyper-parameter tuning. For each model developed, features with near-zero variance were systematically removed. Remaining features were scaled and centered prior to use within AutoML. We used random grid search to tune the hyperparameters. The pipeline includes Gradient Boosting Method, Generalized Linear Model, Random Forest, “Deep Learning” Multi-Layer Perceptrons, and Stacked Ensemble ML (SEML). The list of the hyperparameters optimized for each algorithm in the pipeline is available elsewhere (http://docs.h2o.ai/h2o/latest-stable/h2o-docs/automl.html#faq).

The pipeline generated, optimized, and tested a series of brain age prediction models using the MRI modalities and algorithms listed above. Each model was developed using 5-fold cross-validation, followed by subsequent testing with the held-out testing data set, producing a ranked list of models. Maximum runtime for AutoML was set to 5 hours. *k*-fold cross-validation and the use of a held-out dataset to test model accuracy reduce the risk of over-fitting. Model accuracy was measured using the mean residual deviance (MRD), which is a more useful metric than Mean Squared Error when the distribution is non-Gaussian. We selected the models with the lowest MRD for each modality (sMRI alone and dMRI alone), and their combination (sMRI and dMRI combined), for further assessment. The mean absolute error (MAE), which is measured in years, was also calculated for each model for reference.

### Outcome measures

In addition to the CBCL, we used the following outcome measures: Children’s Global Assessment Scale (CGAS), Flanker Uncorrected Standard Score, and Strengths and Difficulties Questionnaire (SDQ) total difficulties score. The CGAS is a clinically administered assessment of global functioning, with higher scores indicating better functioning. The Flanker task is a measure of sustained attention, with higher scores indicating better performance. The SDQ, like the CBCL, is a general measure of behavioral and emotional symptoms in children, but with fewer questions (25 questions) than the CBCL (118 questions). While there is evidence that the CBCL and SDQ are similar measures (41, 43), there is some evidence that the CBCL better discriminates between community and referred populations (44), and is more sensitive (41), while the SDQ is more specific (41).

### Calculation of Brain Predicted Age Difference (BrainPAD)

We calculated brain predicted age difference (BrainPAD), as described in the literature (24, 45), to capture the difference between the predicted and chronological ages for each participant, with BrainPAD equal to the predicted age minus the chronological age. BrainPAD was therefore positive when the predicted age was greater than the chronological age, reflecting possible neurosupermaturity, and negative when the predicted age was less than the chronological age, reflecting possible neuroimmaturity.

To assess the distinct associations of neurosupermaturity versus neuroimmaturity with the outcome measures, we divided participants based on their BrainPAD values into positive, normal, and negative BrainPAD groups. The positive BrainPAD group was defined by BrainPAD values one standard deviation above the mean; the negative BrainPAD group was defined by BrainPAD values one standard deviation below the mean; and the normal BrainPAD group was defined as a score within one standard deviation from the mean, inclusive.

### Statistical Analyses

Descriptive statistics were generated for the training, testing, and evaluation datasets. We used analysis of variance (ANOVA) tests to evaluate differences in the training, testing, and evaluation groups. Linear models (LM) were used to estimate BrainPAD group averages, assess differences among the BrainPAD groups, and to estimate the association between BrainPAD values and the outcome measures, all while controlling for participants’ age, sex, race, and ethnicity. These analyses were therefore restricted to participants with those data. A separate LM was used for each outcome measure. Plotting and modeling were performed using Rstudio version 1.2.5033 and the R Stats package version 3.6.2 (46). In these analyses, we used the CBCL Total Raw score instead of the Total T-score, as the T-score is already adjusted for age and sex. Throughout, p-values of less than or equal to 0.05 were considered statistically significant.

## Results

### Participant selection and demographics

Workflow for participant selection and analysis is outlined in Figure 1. In brief, of the 585 HBN participants initially eligible for this study, 87 did not have CBCL data and were excluded. The age range among the remaining 498 participants with CBCL data was 5-18. Among the remaining 498 participants, 263 had CBCL Total T-scores less than 60, and 235 had T-scores equal to or greater than 60. Eighty percent of the participants with T-scores less than 60 (N = 215) were included in the training dataset, and 20% (N = 48) in the testing dataset. For the evaluation dataset (N = 249), only participants with race and ethnicity data were included. The evaluation set consisted of 39 participants from the testing set and 210 participants whose CBCL scores were equal to or greater than 60. Demographic information for the participants in the training, testing, and evaluation datasets are displayed in Table 1. Note that while groups differ on clinical markers, the distributions of age, sex, race, and ethnicity are comparable across the three groups.

**Table 1.** Healthy Brain Network Participant Demographics.

|  | **Training Set**<br>**(N = 215)** | **Test Set**<br>**(N= 48)** | **Evaluation Set**<br>**(N = 249)** | **P-value** |
| --- | --- | --- | --- | --- |
| **Age** | 10.79 ($\pm 2.98$) | 10.65 ($\pm 3.42$) | 10.97 ($\pm 3.29$) | 0.74 |
| **Sex** |  |  |  | 0.90 |
| Male | 133 (61.86%) | 29 (60.42%) | 149 (59.84%) |  |
| Female | 82 (38.14%) | 19 (39.58%) | 100 (40.16%) |  |
| **Race** |  |  |  | 0.86 |
| White/Caucasian | 93 (43.26%) | 21 (43.75%) | 127 (51.00%) |  |
| Black/African-American | 34 (15.81%) | 5 (10.42%) | 42 (16.87%) |  |
| Hispanic | 19 (8.84%) | 5 (10.42%) | 23 (9.24%) |  |
| Asian | 6 (2.79%) | 0 (0.00%) | 8 (3.21%) |  |
| Indian | 6 (2.79%) | 1 (2.08%) | 1 (0.40%) |  |
| Native-American Indian | 0 (0.00%) | 0 (0.00%) | 1 (0.40%) |  |
| Two or more races | 32 (14.88%) | 8 (16.67%) | 38 (15.26%) |  |
| Other Race | 4 (1.86%) | 0 (0.00%) | 6 (2.41%) |  |
| Unknown | 2 (0.93%) | 0 (0.00%) | 3 (1.20%) |  |
| Missing | 19 (8.84%) | 8 (16.67%) | 0 (0.00%) |  |
| **Ethnicity** |  |  |  | 0.94 |
| White/Caucasian | 127 (59.07%) | 25 (52.08%) | 167 (67.07%) |  |
| Black/African-American | 48 (22.33%) | 12 (25.00%) | 61 (24.50%) |  |
| Hispanic | 13 (6.05%) | 4 (8.33%) | 15 (6.02%) |  |
| Asian | 4 (1.86%) | 1 (2.08%) | 6 (2.41%) |  |
| Missing | 23 (10.70%) | 6 (12.50%) | 0 (0.00%) |  |
| **CBCL Total Raw Score** | 20.22 | 19.08 | 57.68 ($\pm 25.81$) | < 0.0001 |
| **CGAS Total Score** |  |  |  | < 0.0001 |
| Mean (SD) | 67.91 | 64.74 ($\pm 9.80$) | 61.08 ($\pm 10.19$) | |
| Missing | 116 (53.95%) | 25 (52.08%) | 124 (49.80%) |  |
| **Flanker Uncorrected Standard Score** |  |  |  | 0.46 |
| Mean (SD) | 87.39 | 83.94 | 86.04 ($\pm 15.71$) | |
| Missing | 67 (31.16%) | 16 (33.33%) | 96 (38.55%) |  |
| **SDQ Difficulties Total Score** |  |  |  | < 0.0001 |
| Mean (SD) | 8.81 ($\pm 4.69$) | 8.12 ($\pm 4.57$) | 16.38 ($\pm 6.32$) | |
| Missing | 7 (3.26%) | 0 (0%) | 6 (2.41%) |  |

Between group differences were assessed for the Training Set, Test Set, and Evaluation Set using ANOVA. Notably, no significant differences were found for age, sex, race, or ethnicity.

### Brain age prediction accuracy

Table 2 shows the performance of the most accurate brain age prediction models using morphometry (sMRI), connectomes (dMRI), and combined MRI modalities. Accuracy was determined using the testing dataset. A SEML-based model, applied to the combined MRI modalities, was the most accurate overall (MAE 1.162, MRD 1.983). A SEML-based model was also the most accurate among the tested models using only connectomes. However, with morphometry data alone, the Deep Learning algorithm performed best. Figure 2 shows a scatter plot of the predicted and chronological ages using the SEML-based multimodal model. The ranked list of top 10 tested models for the uni- and multimodal approaches is provided in Supplemental Figures 1a-c.

**Table 2.** Brain Age Prediction Accuracy Across All Models.

| **Model** | **MAE** | **MRD** | **Optimal Algorithm** |
| --- | --- | --- | --- |
| Morphometry and White Matter | 1.1624 | 1.9832 | SEML |
| White Matter Connectomes Alone | 1.3950 | 2.9347 | SEML |
| Morphometry Alone | 1.6530 | 4.5032 | Deep learning (MLP) |

*MAE*: mean absolute error; *MLP*: multi-layer perceptron; *MRD*: mean residual deviance; *SEML*: stacked ensemble machine learning
Table comparing accuracy between unimodal and multimodal models. The highest accuracy was achieved with the model using morphometry and white matter connectomes (sMRI and dMRI combined)

**Figure 2:**
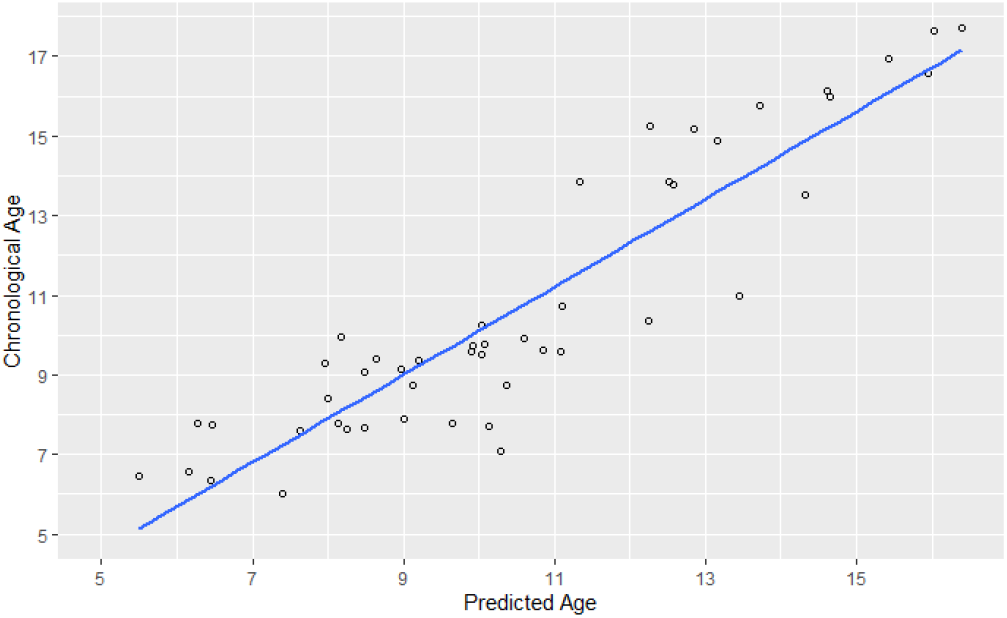
Scatterplot of Predicted Age vs Chronological Age. Scatterplot depicting the relationship between predicted and chronological age for the participants in the held-out test set (N=48) using the morphometry and white matter connectome model.

### BrainPAD, risk of psychopathology, functioning, and cognition

The relationship between brain age, as measured by the most accurate model, and the outcome measures was assessed using the evaluation data set (N = 249). Note that positive BrainPAD values imply neurosupermaturity, while negative scores imply neuroimmaturity. Also, note that higher CBCL scores imply more symptoms, whereas lower CGAS scores imply worse functioning (and typically more symptoms). After controlling for age, sex, race, and ethnicity, lower BrainPAD values were significantly associated with more symptoms on the CBCL (β = −2.325, p = 0.037) and worse functioning on the CGAS (β = 1.850, p = 0.005). Higher BrainPAD values were also significantly associated with better performance on the Flanker (β = 2.372, p = 0.002). There was no apparent association between BrainPAD value and SDQ score (β = −0.197, p = 0.477). CBCL and SDQ scores were correlated in this sample (Pearson’s r = 0.73, p = <0.001).

Scatter plots of BrainPAD and the four outcome measures, along with Loess curves, are plotted in Figure 3. The plots are not adjusted for age, sex, race or ethnicity. The plots are in general agreement with the findings of negative relationships between BrainPAD and CBCL (Fig 3A, top left) and Flanker scores (Fig 3C, bottom left), and a positive relationship with CGAS scores (Fig 3B, top right). A relatively flat line is observed for the relationship between BrainPAD and SDQ score (Fig 3D, bottom left).

**Figure 3:**
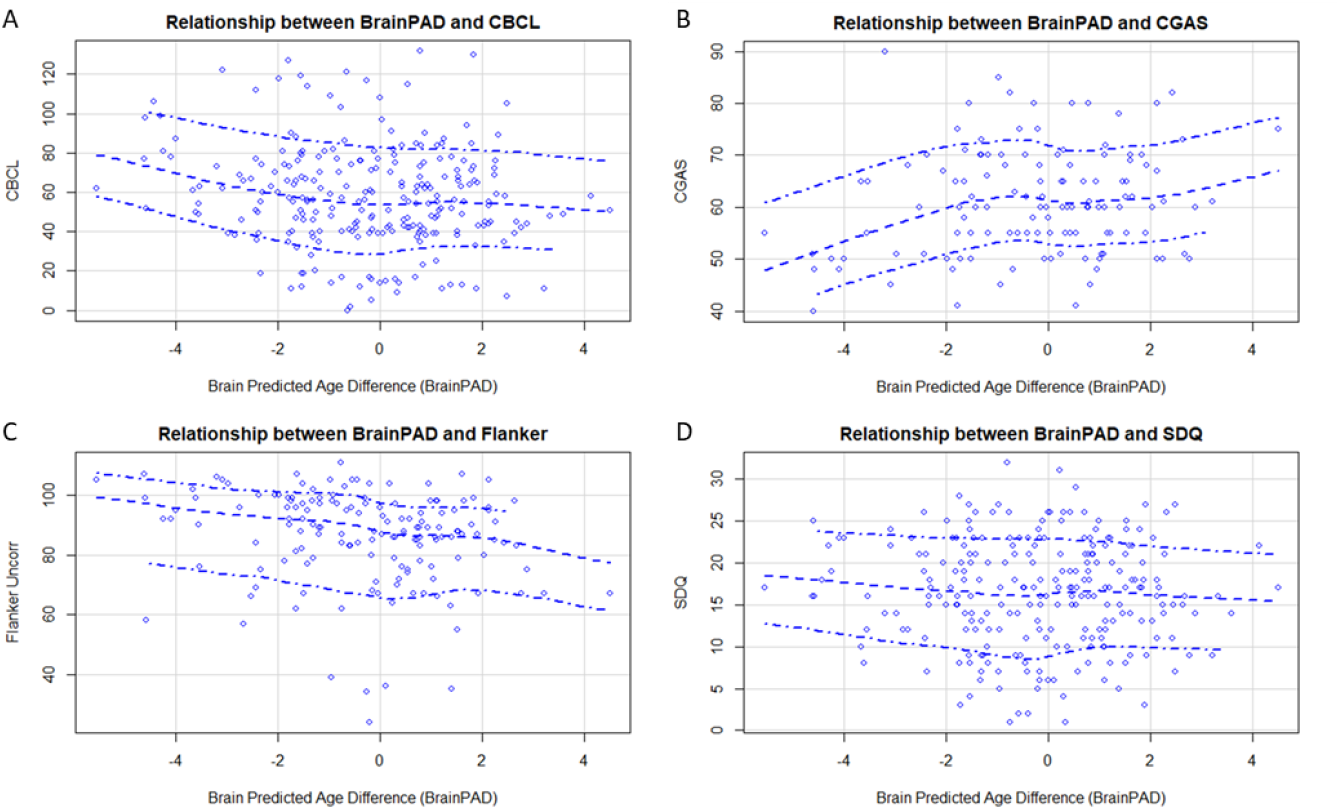
Scatterplot of BrainPAD vs Psychometric Tools. Scatterplots depicting the relationship between BrainPAD and **A**. CBCL, **B**. CGAS, **C**. Flanker, and **D**. SDQ scores among participants in the evaluation dataset (N=249). Loess curves with a 95% confidence interval are shown.

**Figure 4:**
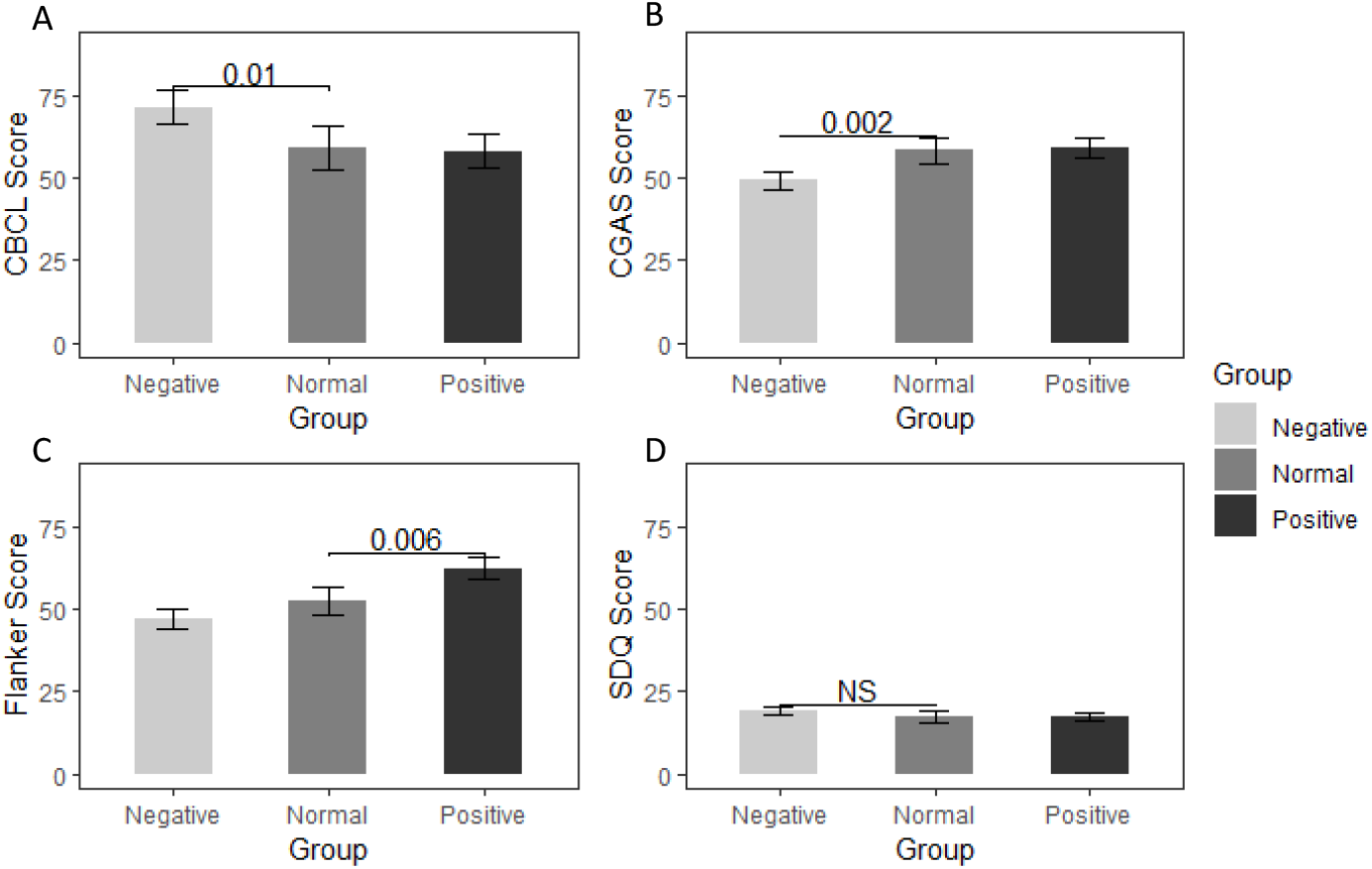
Adjusted Means for 4 Metrics across BrainPAD Groups in the Evaluation Set. Adjusted mean scores for each BrainPAD group in the Evaluation set (N = 249) were calculated for each metric with adjustment for age, sex, race, and ethnicity. A significant difference was noted between the Negative and Normal BrainPAD group for the **A**. CBCL score and **B**. CGAS score. A significant difference was noted between the Positive and Normal BrainPAD group for the **C**. Flanker score only. No significant differences were noted for the **D**. SDQ scores across groups.

We next examined whether the positive, normal, and negative BrainPAD groups differed across our metrics of interest (Figure 3). A statistically significant difference between the negative and normal BrainPAD groups was observed for mean CBCL (negative = 71.6, normal = 59.0, p = 0.011) and CGAS scores (negative = 49.3, normal = 58.3, p = 0.002), but not for the positive and normal groups on these same measures. Additionally, a statistically significant difference was noted between the positive and normal BrainPAD groups for the Flanker task (positive = 62.4, normal = 52.5, p = 0.006), but not for the negative and normal groups (Higher Flanker scores indicate better performance.) There was no statistically significant difference between the normal, negative, or positive BrainPAD groups for the SDQ. The adjusted means for all the outcomes across all three BrainPAD groups is provided in Supplemental Table 2.

## Discussion

In this study, we predicted brain age in youth with and without a high likelihood for psychopathology by applying an automated machine learning pipeline to MRI-derived morphometry and white matter connectomes. We found that morphometry and white matter connectomes together yielded the most accurate brain age prediction, compared to using either modality alone, consistent with prior studies comparing uni- and multimodal approaches (17, 18). Also of note, the SEML method, which combines multiple ML algorithms, was the best predictor of brain age, with an MAE of 1.16 years, suggesting that integrating multiple ML algorithms is advantageous for improving accuracy.

When applying our brain age prediction model to a held-out evaluation dataset, we found associations between deviations from typical brain age, as measured by BrainPAD, and a measure of symptoms (CBCL), functioning (CGAS), and neurocognition (Flanker). Our results indicate that BrainPAD is associated with dysfunction irrespective of reporting source (parents and clinicians) and symptom domain (psychiatric symptoms and neurocognitive performance). We did not find an association between BrainPAD and SDQ scores, despite CBCL and SDQ scores being generally well correlated (41, 43). One possible explanation is that the CBCL better discriminated between participants at high and low risk for psychopathology in this sample. Studies have suggested that the CBCL is a more sensitive measure relative to the SDQ (41), which may explain the more robust relationship between BrainPAD and CBCL than with the SDQ. Taken together, BrainPAD, derived from morphometry and white matter connectomes, may provide a useful objective neuromaturity index that correlates with behavior, functioning, and neurocognition in youth.

Examining the distinct associations for the positive, normal, and negative BrainPAD groups, we found that negative BrainPAD values were associated with more symptoms and poorer functioning. Conversely, participants in the normal and positive BrainPAD groups had comparable symptom burdens and functioning. However, the positive BrainPAD group was distinct from the normal and negative groups in neurocognition, showing superior performance on the Flanker task. This overall pattern appears consistent with the epidemiology of psychiatric disorders linked to neuroimmaturity versus neurosupermaturity. Attentional disorders, which are common overall and particularly in children (47), have been associated with neuroimmaturity (16, 48), and possibly contribute to elevated scores on the CBCL and reduced scores on the CGAS in this age group. Psychotic disorders, which have been associated with neurosupermaturity (14), might also be associated with elevated CBCL and reduced CGAS scores. Even so, psychotic disorders are less common overall and markedly less common in children (49), possibly accounting for the absence of associations between positive BrainPAD and symptoms or functioning. Our finding of a significant association between BrainPAD and Flanker performance is consistent with a previous report linking increased neuromaturity with improved cognitive performance (17). In contrast to studies in adults, neurosupermaturity in this youth sample is associated with better neurocognition, but not with increased risk of psychopathology or poor functioning. Nonetheless, our results do not preclude increased brain age being disadvantageous across other neurocognitive domains, or during other developmental periods (5, 10).

Our study has several limitations. First, as is common to machine learning studies, the resulting model is not easily interpreted, and the use of the SEML algorithm renders the model even less interpretable. Second, our study sample was large enough to develop the model with subsequent testing in a held-out set, but did not have enough participants with specific psychiatric conditions to assess the relationship between neuromaturity and individual DSM-based disorders. Hence, while we detected a general relationship between neuroimmaturity and psychiatric symptoms, and neurosupermaturity and cognitive functioning, we are not able to shed light on the particular neuromaturity-related etiologies of specific disorders, such as schizophrenia (13, 14). Third, this study relies on cross-sectional data, and therefore does not clarify the direction of the relationship between brain changes, symptoms, and functioning.

Fortunately, we see ample opportunities to address these shortcomings in future work. Regarding interpretability, methods are currently in development to specifically enhance the interpretability of the SEML algorithm, such as the use of targeted maximum loss-based estimation (50). As to the sample size, the HBN has continued to collect neuroimaging data. Future studies could deploy the method described in this paper on the updated HBN dataset to establish the replicability of the brain age model and, with sufficient sample size, the relevance of neuromaturity-related etiologies to certain disorders. Finally, the ongoing collection and release of data from the Adolescent Brain and Cognitive Development study, for example, is an opportunity to test the brain age model and explore the association with long-term outcomes on a larger, prospective, longitudinal dataset (51).

In conclusion, our study demonstrates that using multimodal brain imaging, including white matter connectome estimates, and novel, rigorous ML methods, such as SEML, has the potential to improve the accuracy of brain age estimation. Furthermore, BrainPAD shows promise as a general measure of risk for psychopathology and cognitive impairment, with some evidence for distinct associations between particular domains and neuroimmaturity versus neurosupermaturity. Additional work is needed in larger, longitudinal datasets to replicate this approach, clarify causal mechanisms, and make more specific associations between deviations in neuromaturation and specific psychiatric conditions and neurocognitive impairments.

## Data Availability

We used neuroimaging and psychometric data from the HBN collected between 2015, when the study was initiated, through 2017, when HBN neuroimaging data was made publicly available.

http://fcon_1000.projects.nitrc.org/indi/cmi_healthy_brain_network/

## Acknowledgements

We thank the Healthy Brain Network participants and the staff at the Child Mind Institute. Mr. Alex Luna received no financial support for the research, authorship, and/or publication of this article. Dr. Joel Bernanke was supported by Grant No. T32 MH016434-41. Dr. Jiook Cha was supported by Grant No. K01 MH 109836 and by the Brain Pool Program, National Research Foundation of Korea Grant No. 200-20190251. Dr. Jonathan Posner was supported by the Edwin S Webster Foundation, Suzanne Crosby Murphy Endowment under award number R01 MH036197 and UH3 OD023328.

## Supplemental Information

### Supplement

**Table S1a:**
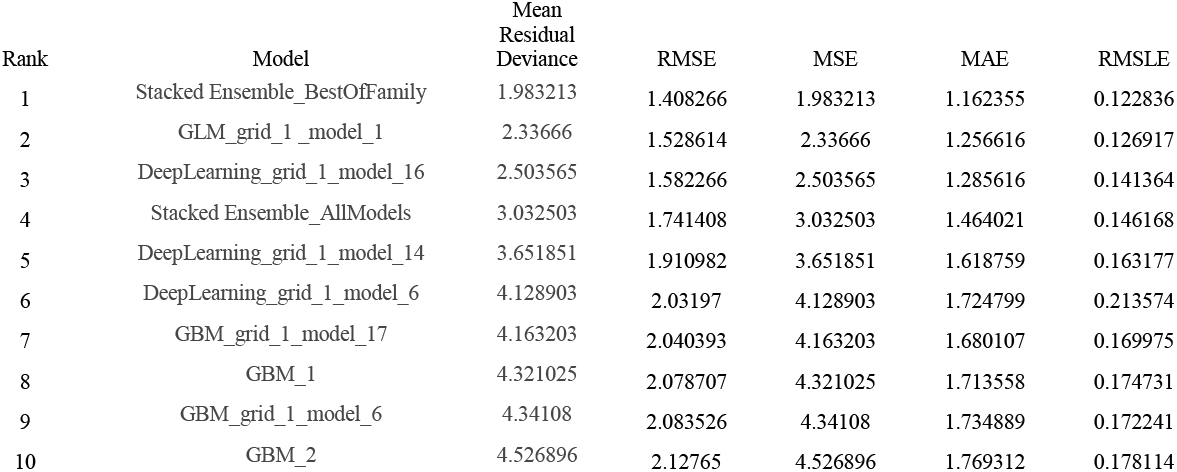
Top 10 Leaderboard – sMRI and dMRI Combined.

**Table S1b:** Top 10 Leaderboard – dMRI Alone.

| Rank | Model | Mean<br>Residual<br>Deviance | RMSE | MSE | MAE | RMSLE |
| --- | --- | --- | --- | --- | --- | --- |
| 1 | StackedEnsemble_BestOfFamily | 2.934726 | 1.713104 | 2.934726 | 1.395017 | 0.150644 |
| 2 | GLM_grid_1_model_1 | 3.17327 | 1.781367 | 3.17327 | 1.436341 | 0.150593 |
| 3 | DeepLearning_grid_1_model_11 | 3.419848 | 1.849283 | 3.419848 | 1.577158 | 0.160439 |
| 4 | DeepLearning_grid_1_model_19 | 3.564923 | 1.8881 | 3.564923 | 1.468145 | 0.151991 |
| 5 | DeepLearning_grid_1_model_6 | 4.474259 | 2.115245 | 4.474259 | 1.778825 | 0.200337 |
| 6 | DeepLearning_grid_1_model_15 | 4.518855 | 2.12576 | 4.518855 | 1.693049 | 0.170429 |
| 7 | GBM_grid_1_model_11 | 4.840511 | 2.200116 | 4.840511 | 1.79599 | 0.183325 |
| 8 | StackedEnsemble_AllModels | 4.849601 | 2.202181 | 4.849601 | 1.816006 | 0.215136 |
| 9 | GBM_grid_1_model_6 | 5.276814 | 2.297132 | 5.276814 | 1.96946 | 0.190546 |
| 10 | GBM_grid_1_model_17 | 5.276979 | 2.297168 | 5.276979 | 1.950674 | 0.188665 |

**Table S1c:** Top 10 Leaderboard – sMRI Alone.

| Rank | Model | Mean<br>Residual<br>Deviance | RMSE | MSE | MAE | RMSLE |
| --- | --- | --- | --- | --- | --- | --- |
| 1 | DeepLearning_grid_1_model_6 | 4.503244 | 2.122085 | 4.503244 | 1.653048 | 0.183928 |
| 2 | DeepLearning_grid_1_model_10 | 4.507287 | 2.123037 | 4.507287 | 1.697663 | 0.182771 |
| 3 | DeepLearning_grid_1_model_9 | 4.532441 | 2.128953 | 4.532441 | 1.650197 | 0.18749 |
| 4 | StackedEnsemble_BestOfFamily | 4.587205 | 2.141776 | 4.587205 | 1.640308 | 0.184302 |
| 5 | DeepLearning_grid_1_model_4 | 4.729697 | 2.174787 | 4.729697 | 1.664533 | 0.188933 |
| 6 | StackedEnsemble_AllModels | 4.731898 | 2.175293 | 4.731898 | 1.708081 | 0.186536 |
| 7 | DeepLearning_grid_1_model_8 | 4.922905 | 2.218762 | 4.922905 | 1.720452 | 0.184681 |
| 8 | DeepLearning_grid_1_model_12 | 5.049262 | 2.247056 | 5.049262 | 1.819278 | 0.188042 |
| 9 | DeepLearning_grid_1_model_2 | 5.138603 | 2.266849 | 5.138603 | 1.866574 | 0.19167 |
| 10 | DeepLearning_grid_1_model_1 | 5.175718 | 2.27502 | 5.175718 | 1.795824 | 0.198884 |

Tables displaying the various accuracy metrics output by H2O AutoML for the multimodal (sMRI + dMRI) or unimodal (sMRI alone or dMRI alone) neuroimaging data. Notably, the Stacked Ensemble models performed best when using sMRI + dMRI and for dMRI alone. As the name implies, Stacked Ensemble – BestOfFamily is based on the best model within each algorithm category (ie Random Forest, GBM, etc), and AllModels is based on every single previously trained model

**Supplemental Table S2:** Adjusted Means For Each Metric Across BrainPAD Group.

|  | CBCL | CGAS | Flanker | SDQ |
| --- | --- | --- | --- | --- |
| Normal BrainPAD | 59.008 | 58.318 | 52.502 | 17.565 |
|  | p = ref | p = ref | p = ref | p = ref |
| Negative BrainPAD | **71.620\*** | **49.279\*\*** | 47.266 | 19.476 |
|  | p = 0.011 | p = 0.002 | p = 0.092 | p = 0.125 |
| Positive BrainPAD | 58.243 | 59.129 | **62.470\*\*** | 17.445 |
|  | p = 0.883 | p = 0.791 | p = 0.006 | p = 0.927 |
| Observations | 249 | 125 | 153 | 243 |

*Note:*
\*p<0.05, \*\*p<0.01, \*\*\*p<0.001
Table displaying adjusted mean scores for each BrainPAD group in the Evaluation set (N = 249) with adjustment for age, sex, race, and ethnicity. Significant differences in the CBCL and CGAS scores between the Negative and Normal BrainPAD groups were observed. A significant difference was noted between the Positive and Normal BrainPAD group for the Flanker score only. No significant differences were noted for the SDQ.

**Table S3:** Correlations between BrainPAD and Various Metrics Within the Test Set.

|  | CBCL | CGAS | Flanker | SDQ |
| --- | --- | --- | --- | --- |
| BrainPAD | -1.674 | 0.703 | 7.189* | -0.279 |
|  | p = 0.325 | p = 0.828 | p = 0.039 | p = 0.708 |
| Age | 0.231 | -0.540 | 4.936*** | 0.228 |
|  | p = 0.737 | p = 0.687 | p = 0.001 | p = 0.454 |
| **Sex (ref = Male)** |  |  |  |  |
| Female | -5.892 | -2.481 | -7.109 | 0.156 |
|  | p = 0.208 | p = 0.736 | p = 0.347 | p = 0.939 |
| **Ethnicity** |  |  |  |  |
| Black/African-American | 3.371 | 4.817 | 3.080 | 2.585 |
|  | p = 0.620 | p = 0.549 | p = 0.741 | p = 0.391 |
| Hispanic | 0.423 | -2.574 | 5.036 | -0.269 |
|  | p = 0.966 | p = 0.805 | p = 0.706 | p = 0.950 |
| Asian | 13.172 | -3.308 | 3.960 | 0.202 |
|  | p = 0.362 | p = 0.830 | p = 0.833 | p = 0.975 |
| **Race** |  |  |  |  |
| Black/African-American | -3.066 | -0.988 | -9.930 | -0.064 |
|  | p = 0.710 | p = 0.922 | p = 0.376 | p = 0.986 |
| Hispanic | -4.978 | -7.573 | 9.669 | -3.307 |
|  | p = 0.589 | p = 0.538 | p = 0.471 | p = 0.417 |
| Indian | -1.105 |  |  | -2.245 |
|  | p = 0.939 |  |  | p = 0.722 |
| Two or more races | -4.391 | 0.256 | -2.365 | -3.046 |
|  | p = 0.455 | p = 0.973 | p = 0.776 | p = 0.243 |
| Constant | 19.696* | 72.577*** | 33.608* | 6.380 |
|  | p = 0.020 | p = 0.001 | p = 0.032 | p = 0.080 |
| Observations | 39 | 20 | 26 | 39 |
| R<sup>2</sup> | 0.199 | 0.204 | 0.591 | 0.100 |

*Note:*
\*p<0.05, \*\*p<0.01, \*\*\*p<0.001
Table displaying the relationship between BrainPAD and various psychometric tools. Of note, the association between CBCL and BrainPAD was similar to that found in the evaluation set ( $\beta = -2.325$ ) but did not reach significance due to insufficient statistical power in the test set ( $N = 39$ )

**Table S4a.** Descriptive Statistics Among CGAS Responders.

|  | **Training Set**<br>**N = 99** | **Test Set**<br>**N = 23** | **Evaluation Set**<br>**N = 125** | **P-value** |
| --- | --- | --- | --- | --- |
| **Age** | 10.42 ($\pm 2.73$) | 10.30 ($\pm 3.07$) | 10.61 ($\pm 3.08$) | 0.84 |
| **Sex** |  |  |  | 0.62 |
| Male | 66 (66.67%) | 17 (73.91%) | 79 (63.20%) |  |
| Female | 33 (33.33%) | 6 (26.09%) | 46 (36.80%) |  |
| **Race** |  |  |  | 0.87 |
| White/Caucasian | 50 (50.51%) | 9 (39.13%) | 67 (53.60%) |  |
| Black/African- | 12 (12.12%) | 4 (17.39%) | 19 (15.20%) |  |
| Hispanic | 9 (9.09%) | 3 (13.04%) | 10 (8.00%) |  |
| Asian | 3 (3.03%) | 0 (0.00%) | 5 (4.00%) |  |
| Indian | 0 (0.00%) | 0 (0.00%) | 0 (0.00%) |  |
| Native-American | 0 (0.00%) | 0 (0.00%) | 1 (0.80%) |  |
| Two or more races | 23 (23.23%) | 5 (21.74%) | 21 (16.80%) |  |
| Other Race | 0 (0.00%) | 0 (0.00%) | 2 (1.60%) |  |
| Unknown | 0 (0.00%) | 0 (0.00%) | 0 (0.00%) |  |
| Missing | 2 (2.02%) | 2 (8.70%) | 0 (0.00%) |  |
| **Ethnicity** |  |  |  | 0.19 |
| White/Caucasian | 59 (59.60%) | 10 (43.48%) | 89 (71.20%) |  |
| Black/African- | 30 (30.30%) | 8 (34.78%) | 27 (21.60%) |  |
| Hispanic | 8 (8.08%) | 2 (8.70%) | 7 (5.60%) |  |
| Asian | 1 (1.01%) | 1 (4.35%) | 2 (1.60%) |  |
| Missing | 1 (1.01%) | 2 (8.70%) | 0 (0.00%) |  |
| **CBCL Total** | 21.40 ($\pm 9.55$) | 20.61 ($\pm 10.65$) | 56.38 ($\pm 24.98$) | < 0.0001 |
| **CGAS** | 67.91 ($\pm 11.18$) | 64.74 ($\pm 9.80$) | 61.08 ($\pm 10.19$) | < 0.0001 |
| **Flanker Uncorrected Standard** |  |  |  | 0.99 |
| Mean (SD) | 87.42 ($\pm 14.14$) | 87.26 ($\pm 14.37$) | 87.66 ($\pm 13.99$) | |
| Missing | 1 (1.01%) | 0 (0%) | 0 (0%) |  |
| **SDQ Difficulties** |  |  |  | < 0.0001 |
| Mean (SD) | 8.89 ($\pm 4.26$) | 8.09 ($\pm 3.81$) | 15.74 ($\pm 6.24$) | |
| Missing | 1 (1.01%) | 0 (0%) | 0 (0%) |  |

Table displaying descriptive statistics among participants who participated in the CGAS questionnaire. Between group differences were assessed for the Training Set, Test Set, and Evaluation Set using ANOVA. Notably, no significant differences were found for age, sex, race, or ethnicity.

**Table S4b.** Descriptive Statistics Among Flanker Responders.

|  | **Training Set**<br>**N = 148** | **Test Set**<br>**N = 32** | **Evaluation Set**<br>**N = 153** | **P-value** |
| --- | --- | --- | --- | --- |
| **Age** | 10.57 ($\pm 2.87$) | 10.51 ($\pm 3.37$) | 10.80 ($\pm 3.27$) | 0.77 |
| **Sex** |  |  |  | 0.87 |
| Male | 93 (62.84%) | 20 (62.50%) | 92 (60.13%) |  |
| Female | 55 (37.16%) | 12 (37.50%) | 61 (39.87%) |  |
| **Race** |  |  |  | 0.81 |
| White/Caucasian | 59 (39.86%) | 11 (34.38%) | 80 (52.29%) |  |
| Black/African-American | 25 (16.89%) | 5 (15.62%) | 24 (15.69%) |  |
| Hispanic | 14 (9.46%) | 4 (12.50%) | 12 (7.84%) |  |
| Asian | 5 (3.38%) | 0 (0.00%) | 7 (4.58%) |  |
| Indian | 2 (1.35%) | 0 (0.00%) | 0 (0.00%) |  |
| Native-American Indian | 0 (0.00%) | 0 (0.00%) | 1 (0.65%) |  |
| Two or more races | 27 (18.24%) | 7 (21.88%) | 27 (17.65%) |  |
| Other Race | 3 (2.03%) | 0 (0.00%) | 2 (1.31%) |  |
| Unknown | 0 (0.00%) | 0 (0.00%) | 0 (0.00%) |  |
| Missing | 13 (8.78%) | 5 (15.62%) | 0 (0.00%) |  |
| **Ethnicity** |  |  |  | 0.18 |
| White/Caucasian | 79 (53.38%) | 15 (46.88%) | 110 (71.90%) |  |
| Black/African-American | 39 (26.35%) | 9 (28.12%) | 33 (21.57%) |  |
| Hispanic | 12 (8.11%) | 3 (9.38%) | 7 (4.58%) |  |
| Asian | 3 (2.03%) | 1 (3.12%) | 3 (1.96%) |  |
| Missing | 15 (10.14%) | 4 (12.50%) | 0 (0.00%) |  |
| **CBCL Total** | 20.84 ($\pm 9.77$) | 18.88 ($\pm 10.96$) | 56.82 ($\pm 26.35$) | < 0.0001 |
| **CGAS** |  |  |  | < 0.0001 |
| Mean (SD) | 67.90 ($\pm 11.23$) | 64.74 ($\pm 9.80$) | 61.08 ($\pm 10.19$) | |
| Missing | 50 (33.78%) | 9 (28.12%) | 28 (18.30%) |  |
| **Flanker Uncorrected** | 87.39 ($\pm 13.91$) | 83.94 ($\pm 17.89$) | 86.04 ($\pm 15.71$) | 0.46 |
| **SDQ Difficulties** |  |  |  | < 0.0001 |
| Mean (SD) | 9.08 ($\pm 4.48$) | 7.44 ($\pm 4.06$) | 15.63 ($\pm 6.36$) | |
| Missing | 2 (1.35%) | 0 (0%) | 0 (0%) |  |

Table displaying descriptive statistics among participants who participated in the Flanker task. Between group differences were assessed for the Training Set, Test Set, and Evaluation Set using ANOVA. Notably, no significant differences were found for age, sex, race, or ethnicity.

**Table S4c.** Descriptive Statistics Among SDQ Responders.

|  | **Training Set**<br>**N = 208** | **Test Set**<br>**N = 48** | **Evaluation Set**<br>**N = 243** | **P-value** |
| --- | --- | --- | --- | --- |
| **Age** | 10.71 ($\pm 2.95$) | 10.65 ($\pm 3.42$) | 10.94 ($\pm 3.27$) | 0.68 |
| **Sex** |  |  |  | 0.91 |
| Male | 129 (62.02%) | 29 (60.42%) | 146 (60.08%) |  |
| Female | 79 (37.98%) | 19 (39.58%) | 97 (39.92%) |  |
| **Race** |  |  |  | 0.87 |
| White/Caucasian | 92 (44.23%) | 21 (43.75%) | 126 (51.85%) |  |
| Black/African- | 31 (14.90%) | 5 (10.42%) | 40 (16.46%) |  |
| Hispanic | 18 (8.65%) | 5 (10.42%) | 22 (9.05%) |  |
| Asian | 6 (2.88%) | 0 (0.00%) | 7 (2.88%) |  |
| Indian | 6 (2.88%) | 1 (2.08%) | 1 (0.41%) |  |
| Native-American | 0 (0.00%) | 0 (0.00%) | 1 (0.41%) |  |
| Two or more races | 31 (14.90%) | 8 (16.67%) | 37 (15.23%) |  |
| Other Race | 4 (1.92%) | 0 (0.00%) | 6 (2.47%) |  |
| Unknown | 2 (0.96%) | 0 (0.00%) | 3 (1.23%) |  |
| Missing | 18 (8.65%) | 8 (16.67%) | 0 (0.00%) |  |
| **Ethnicity** |  |  |  | 0.91 |
| White/Caucasian | 124 (59.62%) | 25 (52.08%) | 165 (67.90%) |  |
| Black/African- | 46 (22.12%) | 12 (25.00%) | 58 (23.87%) |  |
| Hispanic | 13 (6.25%) | 4 (8.33%) | 14 (5.76%) |  |
| Asian | 4 (1.92%) | 1 (2.08%) | 6 (2.47%) |  |
| Missing | 21 (10.10%) | 6 (12.50%) | 0 (0.00%) |  |
| **CBCL Total** | 20.21 ($\pm 10.10$) | 19.08 ($\pm 10.91$) | 57.35 ($\pm 25.64$) | < 0.0001 |
| **CGAS** |  |  |  | < 0.0001 |
| Mean (SD) | 67.94 ($\pm 11.23$) | 64.74 ($\pm 9.80$) | 61.08 ($\pm 10.19$) | |
| Missing | 110 (52.88%) | 25 (52.08%) | 118 (48.56%) |  |
| **Flanker Uncorrected Standard** |  |  |  | 0.53 |
| Mean (SD) | 87.14 ($\pm 13.84$) | 83.94 ($\pm 17.89$) | 86.04 ($\pm 15.71$) | |
| Missing | 62 (29.81%) | 16 (33.33%) | 90 (37.04%) |  |
| **SDQ Difficulties** | 8.81 ($\pm 4.69$) | 8.12 ($\pm 4.57$) | 16.38 ($\pm 6.32$) | < 0.0001 |

Table displaying descriptive statistics among participants who participated in the Strengths and Difficulties questionnaire. Between group differences were assessed for the Training Set, Test Set, and Evaluation Set using ANOVA. Notably, no significant differences were found for age, sex, race, or ethnicity.

